# Considering causality in normal tissue complication probability model development: a literature review

**DOI:** 10.1101/2025.07.09.25331182

**Authors:** Alexa M.M. Mulder, Jungyeon Choi, Lotta M. Meijerink, Wouter van Amsterdam, Artuur M. Leeuwenberg, Ewoud Schuit

## Abstract

**Background:** normal-tissue probability models that estimate the probability of complications often associated with radiotherapy could potentially be used to help clinicians make decisions regarding the radiation dose or the type of radiation treatment. In order to be able to use NTCP models in this way, they should accurately capture the causal relation between dose and complication risk. The question then remains: do current normal-tissue complication probability models for radiation treatment optimization look at causality during model development?

**Objective:** to evaluate the consistency of causal statements in existing NTCP model development studies discussing the relationship between radiation and complications in patients with head and neck cancer. *Methods*: a comprehensive search by a recent systematic Cochrane review was used to obtain articles reporting on the development and external validation of NTCP models that predicted complications. The full text of all the relevant articles were assessed for: stated aim; claims for potential use; if adjustments for confounding were made; and use of language implying causality.

**Results:** out of the 98 evaluated studies, the minority (11.2%) stated causal aims even though 43.9% of studies made causal recommendations. Overall, 31.6% studies started out with an apparent predictive intent but ended up making causal claims in their conclusion or discussion. Out of all the studies that made causal recommendations there were none that explicitly adjusted for confounding.

**Conclusion:** misalignment between the aims of studies and the interpretation of their results in term of causality is common in observational research of NTCP models for patients with head and neck cancer. Researchers should precisely express their aim; if their aim is to make causal recommendations, they should at least discuss and consider confounding factors when formulating their study design.

## Introduction

While radiation therapy has been shown to improve long term survival in patients with head and neck, it may induce various complications as a result of radiation exposure to surrounding normal tissues. These complications include xerostomia, dysphagia, and hypothyroidism, all of which can have a significant impact on quality of life. It is therefore important to try to maintain the required dose to the malignant tissue while simultaneously trying to prevent radiation-induced toxicity.

Normal Tissue Complication Probability models (NTCPs) describe the relationship between dose distributions in organs at risk (OAR) and the development of complications. These models are developed for the purpose of estimating the risk of radiation-induced complications. In clinical practice, they may inform patients and physicians of the probability of a future outcome, which can then be used to inform patients or provide decision support.

Randomized controlled trials are regarded the gold standard for studying causal relationships to base treatment decisions on.^1^ However, randomizing individuals is often not feasible or appropriate, and there has been a growing interest towards undertaking causal inference from observational data.^2^ NTCP models based on observational data may then be used to optimize dosage plans and to support the decision on what type of radiation treatment to give.^3^ This has already been introduced in practice, for example, in the Netherlands, a model-based approach is used in which selection of patients is based on a predicted clinically relevant reduction of complications.^4^ However, if NTCP models are to be used as a basis to intervene on the predictor values (i.e. dosimetric predictors), these models need to need to accurately capture the causal relation between dose and the risk of side effects.^5^

Both studies with a causal intent as well as studies with a purely predictive intent can be performed on the same observational data, but the underlying research question, methods and interpretation of results should be adapted to their respective purpose.^6^ To be able to make any claims about the potential usage of their models for applications that assume causality, such as dose optimisation or plan comparison, the study needs to consider confounding factors. ^7^

In this context, confounders influence the development of complications as well as the radiation dose. For example, when investigating a causal relationship between radiation dose and development of complications, tumour location can be a confounding factor. The tumour location may contribute to complications; a tumour located in the parotid gland may increase the likelihood of a patient suffering from xerostomia. In addition, radiologists determine the radiation dose and target area based on the location of the tumour and its relation to important surrounding structures.^8^ Other examples of potentially confounding factors include patient-related factors (i.e., gender, presence of complications at baseline, comorbidities, genetic factors, medication), disease-related factors (i.e., tumour location), and treatment-related factors (i.e., type of radiation treatment, use of chemotherapy). If confounding is not properly considered, it can result in a distortion of the true association and therefore may lead to wrong conclusions regarding the radiation’s impact on the outcome of interest.^9^

The most common way to reduce confounding bias is by using various types of statistical analysis.^10^ Studies may measure and report potential confounders and use randomisation, restriction/matching, or inclusion in multivariate models to reduce confounding bias. It is important to note that randomization reduces both measured and unmeasured confounding bias, while restriction and matching adjustments in multivariate models only addresses measured confounding bias. When studies use multiple variables in their models, it is also of importance to think carefully about how the selection of these variables is made. Automatic variable selection may work for purely predictive models but not necessarily for causal inference since these may not always select all known confounders.^11^ Finally, these studies may choose to include covariates in a multivariate model for other reasons than to adjust for confounding. This is significant because when not used for the right reasons, these methods might not be implemented correctly.^11^

Results from prediction research cannot be interpreted causally without considering methodological issues in causal research such as confounding bias. This is why it would be of interest to review existing NTCP model development studies in the literature and assess to what degree the research question, the model development methodology, and the claims concerning results capture the causal dose-complication relation.

In this study we aim to assess existing studies discussing NTCP models that predict complications that could be caused by radiotherapy in order to elucidate any statement they make regarding causality, either in their stated goal or in the claims they make for the use of the model. Additionally, this study will evaluate the methodology of these studies for any potential adjustments that are made for confounding factors.

## Methods

### Description of included articles and search strategy

A comprehensive search using Ovid MEDLINE, Ovid Embase, ClinicalTrials.gov and World Health Organization International Clinical Trials Registry Platform was performed. All searches were run in March 2021. The retrieved articles were screened for additional references. Articles were included that reported on the development and external validation of NTCP models that are designed to predict complications in patients with head and neck cancer after radiation treatment. The full search strategy can be found in the protocol by Takada et al. and the abstract by Tambas et al.^12,13^

### Data extraction and assessment

The following information was extracted after the full-text screening:

#### General information

- Title
- Main author
- Year of publication
- Predictors considered for the model
- Predictors included in the model
- Notes on the choice of predictors
- Outcome (i.e., complications)

#### Aim of the study

- We classified study intent as causal, non-causal or unclear based on whether the stated aim of the study is to make a model that is only intended for prediction of the outcome or to make a model that could be used to make clinical decisions regarding the dose or type of radiation therapy. Studies were considered to be unclear in their intent if authors used ambiguous terminology that may be considered causal when stating aims. This classification was done based on sentences extracted verbatim from the abstract or introduction starting with phrases like: “The aim of this study was…” or “The objective of this article is…”.
- Additional statements relating to the aim of the study.

#### Statements regarding the potential use of the study

- We evaluated any claims the authors made about the potential use of the model as stated in their discussion or conclusion. Statements were considered causal if the study mentioned dose adjustments that could be made based on their models or if studies gave recommendations regarding the type of radiation treatment that would be most suitable. Studies that made treatment recommendations based on prediction were not considered casual. For example, if the model can be used to identify high risk patient in order to start treatment to minimise side effects early. Statements were considered “unclear” if the studies made use of phrases and language that could be interpreted as causal. Causal language is defined as language used to indicate situations in which the exposure directly influences the outcome. Decisions on whether words or phrases could be considered causal were based on the interpretation of the reviewer using the guidelines created by the AMA (see appendix 1).^14,15^

#### Methods for confounding adjustment

- Any methods used that could be potentially used as adjustments for confounding. These include using matching to aim for equal distribution of confounders; restricting entry to the study of individuals with confounding factors; or using multivariate analysis.
- Method of covariate selection.
- Whether these methods were explicitly used for the purpose of adjusting for confounding. This is an important criteria because for the adjustments to be effective, researchers have to think carefully about the selection of potential confounders because the effect of adjustments for confounding is naturally limited to known (and measured) confounding factors.^16,17^
- Additional statements relating to the claims of the studies, especially whether there was any theoretical discussion about causal relationships between the exposure and outcome and whether explicit causal disclaimer statements (e.g., “this study could not make any causal recommendations”).

The data extraction was done by one independent reviewer. Any statements regarding purpose or claims the reviewer was not sure on how to classify, was discussed with two other researchers until consensus was reached.

## Results

A total of 99 unique potentially relevant articles were identified. One was excluded because it was only available in French. The remaining 98 articles were used to extract data. The year of publication ranged from 1999 to 2021.

### Aim/purpose of the study

Non-causal aims were the most common (80 studies, 81.6%), followed by causal aims (11 studies, 11.2%). The remaining 7 studies (7.1%) were not clear on their definite purpose but made statements that could be interpreted as both causal or non-causal. Table 1 shows sample excerpts of phrases used to describe the aim of the study. Words referring to the particular topic of the corresponding study were removed from the statements.

**Table 1:**
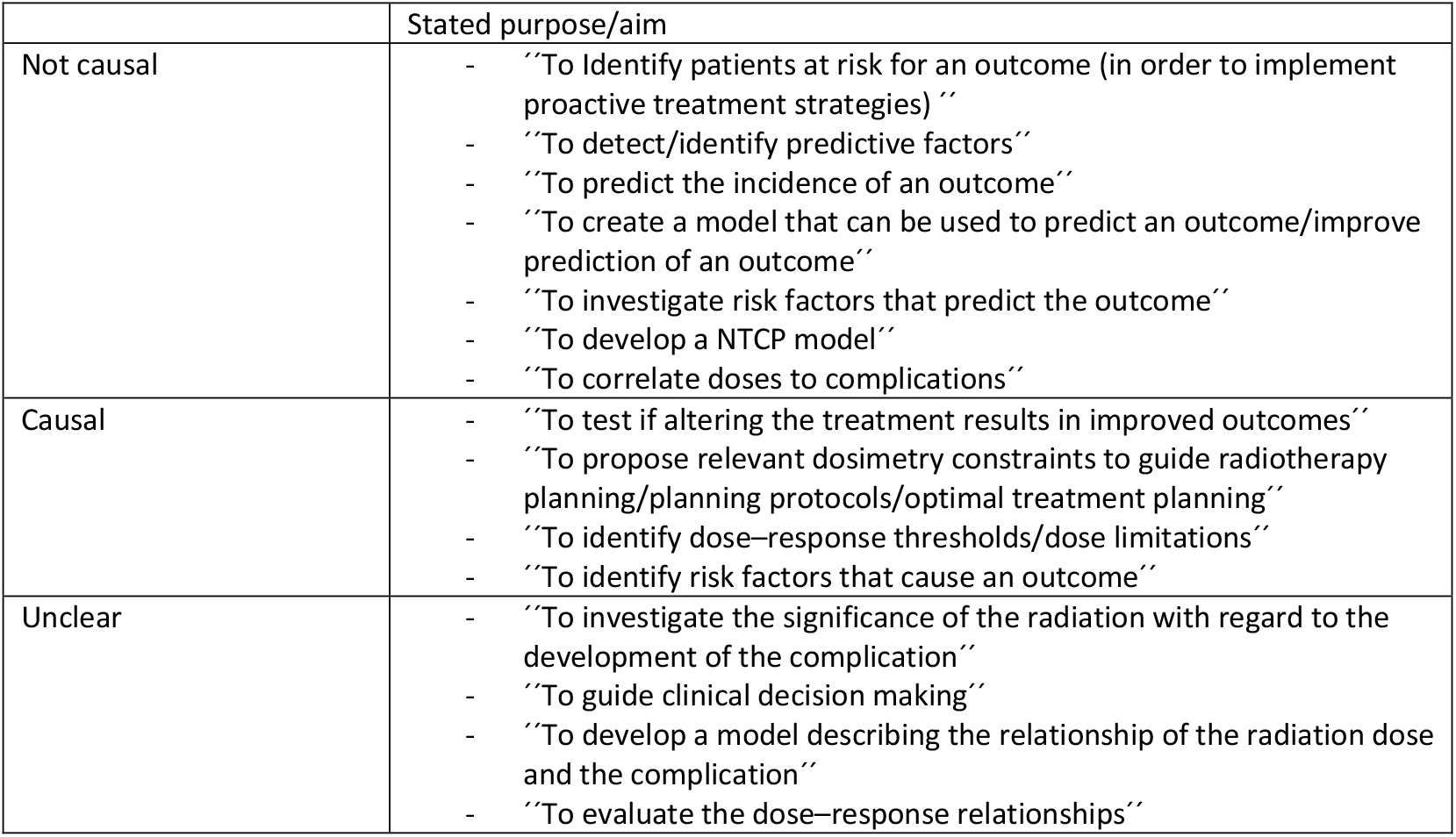
examples of the stated purpose or aim of different studies.

### Conclusions and recommendations

There were 45 (45.9%) studies that made clear causal recommendations, and 42 (42.9%) that refrained from making any causal statements in their discussion or conclusion. Studies were classified as causal when they made clear statements regarding dose adjustments. Studies were classified as having no causal claims if they intended their model only to be used as a predicove tool. Table 2 shows phrases that implied a clear causal relaoonship between dose adjustment and radiaoon-induced toxicity.

**Table 2:**
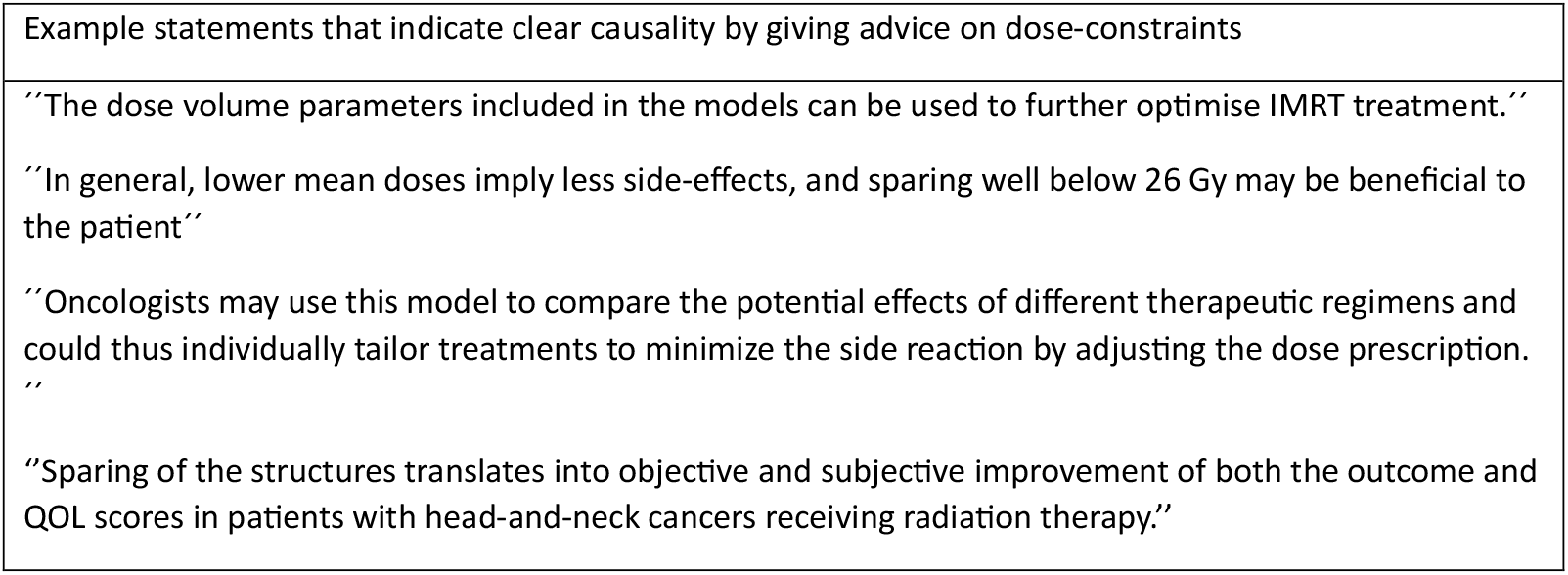
example statements extracted from the discussion or conclusion of different studies, indicating either a causal purpose or a non-causal purpose.

The remaining 11 (11.2%) studies were classified as unclear. These studies did not explicitly suggest dose-adjustment based on their study results but implied a causal relaoonship between dose and outcome. Table 3 shows the phases and wording that could infer causality.

**Table 3:** studies that were classified as “unclear” and the language that was used that might imply causality.

| Study name | Relevant statements extracted from the conclusion or discussion | Comments |
| --- | --- | --- |
| Alterio (2017) | “Our results suggested that the absorbed dose **might play a relevant role** in the development of the outcome.” | “Playing a relevant role” could be interpreted as the radiation influencing the outcome. |
| Cheraghi (2017) | “The results showed that some clinical parameters **have great impacts on** radiotherapy induced SNHL (p value <.05).” | The use of the word “impact” implies that the claim is causal. |
| Dijkema (2010) | “ **When aiming at** preservation of parotid gland function after RT for HN tumors, this study shows a gradual increase in NTCP with increasing mean dose. In fact, a treatment planning constraint of 25–30 Gy **corresponds to** 17–26% complication probability at 1 year.” | This implies that this data can be used to prevent the outcome by lowering the dose. However, the study does not make any explicit recommendations. |
| Pota (2017) | “Predictors are detected, and **their influence on each outcome is revealed.**” | The influence on an outcome could be interpreted as causal. |
| Sheikh (2019) | “Taken together, prediction models based on these features **can further our understanding of the development of** radiation-induced xerostomia and allow us to develop patient-specific adaptations to radiation treatment plans to minimize toxicity.” | Understanding the development of the complications suggests a causal relationship. |
| Soares (2018) | “The role of age, gender, severity of xerostomia prior to radiation therapy and planned mean physical dose in the contralateral and ipsilateral parotids appeared to be of **main importance to the development of the radiation** therapy side-effect xerostomia.” |  |

There were seven studies that made causal claims, but added the caveat that these recommendations should only be followed after future research has been done. We classified these as making “no causal claims”.

Seven studies mentioned in their conclusion that their study results could be used for treatment planning; for example, using phrases like: “This model could be used for: optimising treatment, individualising treatment, designing a more suitable treatment plan etc.”. For these studies, we could not judge whether the authors implied or predictive interpretation of the study results. These could be interpreted as giving recommendations on dose optimisation. However, it could also be read as implementing preventative measures. For instance, a study might have identified high risk patients for severe weight loss after radiation treatment based on the NTCP model developed. One may use this information to personalize the dose-adjustment for the high-risk patients in order to reduce the weight loss, which implies a causal interpretation of the model. Another person may prescribe a prophylactic feeding tube for patients who are at high risk of severe weight loss after radiation treatment, which implies a predictive interpretation of the model. In this study, we labelled these studies as “unclear” in their claims.

Table 4 shows a summary of all of the included studies and whether their intent or claims was classified as causal, non-causal or unclear (the full table with all of the studies and their classification can be found in appendix 2). There were 49 (50.0%) studies that were consistent with their purpose and claims; they were either both causal or they were both non-causal. One study was unclear, both in its purpose and its claims. The remaining 48 (49.0%) were misaligned in their intent and claims. Overall, there were 32 (32.7%) studies started out with an apparent predictive intent but ended up making causal claims in their conclusion or discussion.

**Table 4:**
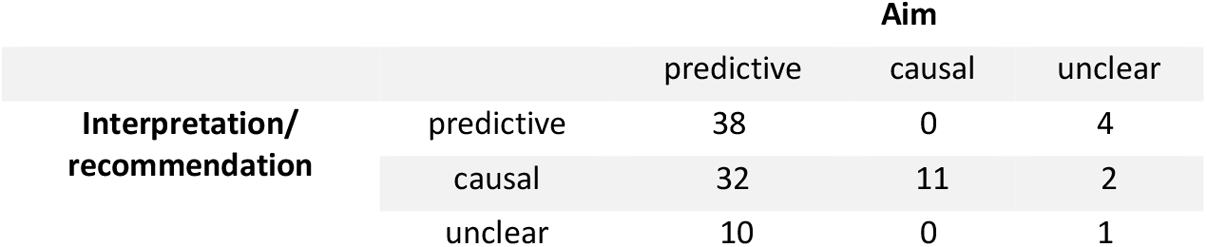
overview of the classification of the aim and interpretation/recommendation of the studies evaluated in this article.

|  |  | Aim |  |  |
| --- | --- | --- | --- | --- |
|  |  | predictive | causal | unclear |
| Interpretation/<br>recommendation | predictive | 38 | 0 | 4 |
|  | causal | 32 | 11 | 2 |
|  | unclear | 10 | 0 | 1 |

### Adjustments for confounding

Studies with clear causal claim had several ways of selecong the predictors for their final models. Some of these methods could be used to reduce confounding. We have classified these methods as follows:

1.The use of restriction. These studies only included certain subjects that have the same values of potential confounding variables. This eliminates variation in the confounder. Two types of restriction were used. First, exclusion of patients who had baseline dysfunction of the outcome. Second, the use of any medication known to affect the outcome was prohibited, or none of the patients included in the study used certain medications that could be expected to change the dose-response relationship.

2.Using statistical methods. Either studies made use of univariate analysis to ascertain whether certain non-dosimetric predictors were significantly associated with the outcome or they inserted several non-dosimetric variables into a multivariate model. Studies were not considered to have used statistical adjustments if they only use dosimetric parameters in their multivariate models.

3.If the study used statistical adjustments, and therefore included non-dosimetric variables in their model, we assessed if there was any mention in the text on how these variables were selected. Some studies based their predictors on prior knowledge, literature, or clinical expertise, while some models just used predictors that were available to them from a database without any clear substantiation.

Table 5 presents these methods that are used by studies that make causal recommendations, the number of studies that employed them and whether the study explicitly stated if these methods were used to account for confounding factors. The full details on each separate study can be found in appendix 3.

**Table 5:**
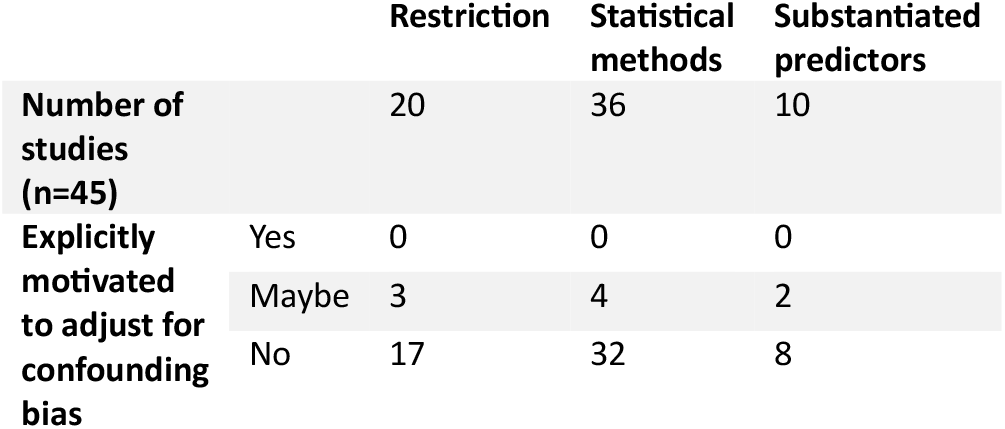
number of studies with a clear causal aim (n=45) and the methods that they could have potentially used to adjust for confounding.

Out of the studies that made causal recommendations, there were seven that did make any adjustments for confounding factors.

There were ten studies that acknowledged that there are factors that could potentially affect the outcomes that could not be accounted for in their study. There were only four studies that mentioned somewhere in the text why they employed these specific methods. None of them explicitly reported that these were used to adjust for confounding and thus are labelled as “maybe” in table 5. Beetz et al. stated the following: “Xerostomia is a complex endpoint, which can be influenced by several. Although we tried to take these factors into account as much as possible, there will be other prognostic factors that remained unidentified”. Chao et al. and Lee at al. were more specific in their statements. Stating, respectively: “To examine whether the correlation between outcome and irradiation dose depends on other potential contributing factors, we performed a multivariate analysis” and “To ensure that xerostomia is induced primarily by radiation treatment patients with baseline dysfunction were excluded from analysis”. The remaining studies did not mention anywhere in the text for what exact purpose the methods of restriction or statistical analysis were used.

### The use of the word “confounding”

There were six studies that specifically mentioned “confounders” or “confounding”. In only three of these studies, it was used in the true sense of the word. Namely, a factor that is predictive of the outcome, associated with the exposure, and is not an intermediary between the exposure and the outcome. The first one cited concomitant chemotherapy, fractionation schedules and the primary tumour sites as confounders. The second one stated the mean dose as a confounder (their primary goal was to find a relationship between the presence of genes that are associated with radiosensitivity and the outcome). The third used the word broadly: “Although interfering factors were excluded as much as possible, there is an inevitability of confounding factors associated with the retrospective analysis”. In the remaining three studies that mentioned the word confounding, the text was unclear either to what variable the word referred to, or to what the association of the variable referred to had with either the association or the outcome.

## Discussion

### Main findings

This review evaluated how studies focussed on the development of NTCP models using observational data communicated research aims and conclusions, focussing specifically on the role of causality. For the studies that made causal recommendations, we investigated if or how they accounted for confounding. The most notable finding was that 32.7% of studies made causal claims in a way that did not match the stated non-causal intent of their research. Out of the studies in this paper that made causal claims, there were none that explicitly adjusted for confounding. This would imply that the majority of the studies that make causal claims do not make adequate adjustments to substantiate their conclusions. Finally, 19.4% of studies formulated either an unclear purpose, or made unclear interpretations of their results due to ambiguous use of possible causal language.

Our findings imply that there is often a mismatch between research aim (predictive vs. causal) and the interpretation of the results in NTCP research. It has been previously found that authors may be eager to make causal claims when interpreting their results. This is possibly because causal inference is the ultimate goal of most clinical and public health research.^18^ Gleadhill et al found that non-causal aims are most common, which corresponds to our findings.^19^ The discrepancy between having a non-causal purpose and making causal claims was one of the more noteworthy findings in our study. This could not only cause interpretability issues for the reader; it could also mean that studies are less likely to adjust for bias when creating the methodology.

Most studies that claimed causal interpretation of their results did not correctly account for confounding. There are several papers that have been published on how to conduct etiological research on observational data. If the methodology considers bias that can be caused by confounding, it is possible to make causal claims using observational data. When making causal inference, it is vital to think critically about what variables can be confounders based on previous literature and knowledge of the subject matter.^3^ Automatic variable selection or variable selection based solely on the data available in the database may work when creating prediction models, but not necessarily for making causal claims.^20^

Finally, our study revealed that many articles used unclear or ambiguous causal language. Olarte et al. found that potential causal words and phrases are embedded in observational research, regardless the purpose of that research.^21^ Even though only 19.4% of articles in our study were unclear in formulating the aim of the study or in interpreting the results, this is still a significant number. Published research, especially articles that might be used to influence clinical decision-making, has important knowledge implications and it is therefore crucial for authors to use language that is clear and appropriate to their work. Use of unclear causal language can cause misinterpretation of the text, not only by scientifically trained readers but especially by media reporters, clinicians, and the general public. This misinterpretation can lead to potentially serious consequences, including erroneous medical decisions.^22^

### Recommendations

We would recommend authors to clearly specify their intent, whether it is causal or predictive, when formulating a research question. Once the aim is clear, we would recommend consulting appropriate methodological guidelines as well as methodological experts.^23,24^ Finally, we would suggest using language throughout the article that is consistent with the intent of the study.

### Strengths and limitations

The current study has a number of strengths. We performed full text screening of all currently available articles discussing the development of NTCP models for patients with head and neck cancer using a comprehensive search strategy. Any doubts on classifying statements were discussed with two other are experienced researchers and statisticians. Limitations include the fact that he data-extraction itself was performed by a single researcher. The evaluation will invariably contain some subjectivity. It has been shown that the interpretation of causal language can vary widely between reviewers.^25^ Furthermore, some of the included studies were written by authors whose first language was not English, which could cause misinterpretation of their intent. Our analysis was limited to the field of head and neck cancer and results might be different in other fields. To amend this, we also looked at two systematic reviews in the field of lung cancer. These studies showed similar outcomes to the ones shown in the result section. The full evaluation of these articles can be found in appendix 4. Finally, we did not consider the quality of the studies included in our assessment.

## Conclusion

When developing NTCP models that estimate the probability of complications after radiotherapy, there are many researchers that remain tempted to draw causal conclusions from observational data while the purpose and methods were not focussed on causality. These models will not accurately capture the causal relation between dose and complication if they do not adequately adjust for confounding. It is important to determine causal factors correctly if these models are to be used as an incentive to change clinical patient care. We recommend that authors should openly discuss the possible discrepancies between the data-based measure they are able to estimate and the desired targeted causal effect. To achieve this purpose, researchers should be explicit about their aims. If causal inference is the end-goal, researchers should make sure potential confounding factors are adjusted for and whether identification of such factors was plausible and comprehensive.

## Data Availability

All data produced in the present work are contained in the manuscript.

---

**Appendix 1:** terminology of causation.

| Causal language | Non-causal language |
| --- | --- |
| Causes<br>Effects, modifies<br>Makes<br>Influences<br>Impacts<br>Results in<br>Induces<br>Effective in<br>Is attributable to, contributes to<br>Leads to<br>Responsible for | Associated<br>Related<br>Correlated<br>Predicts<br>Linked to<br>Varies with |

**Appendix 2:**
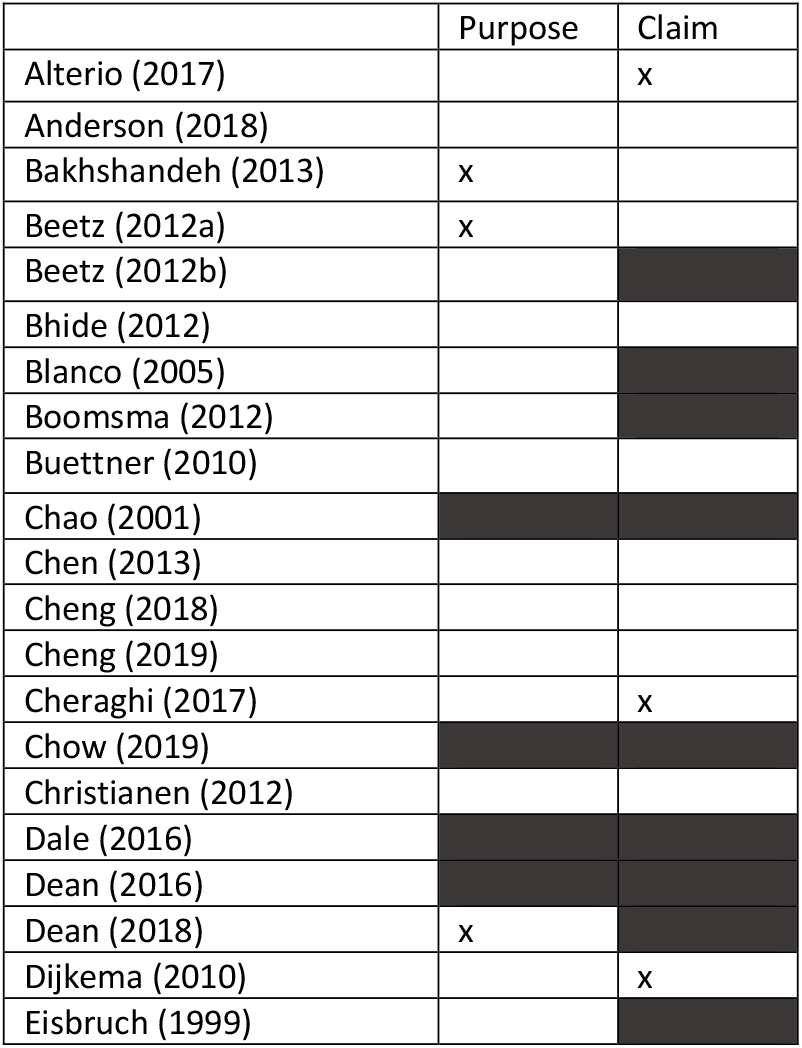
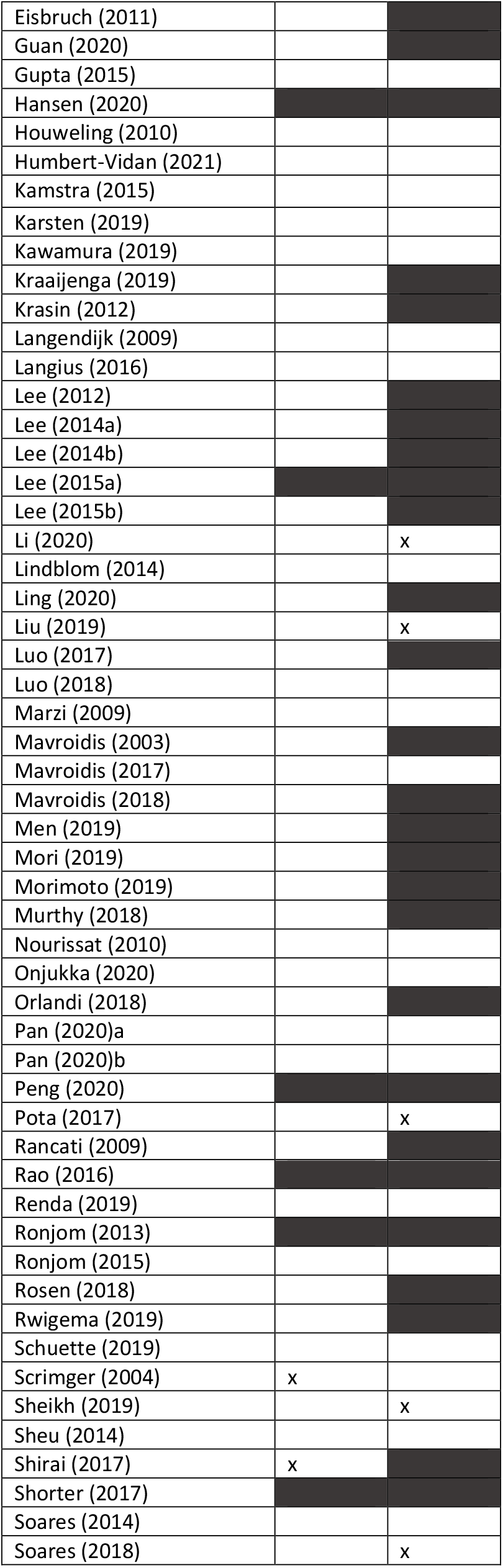
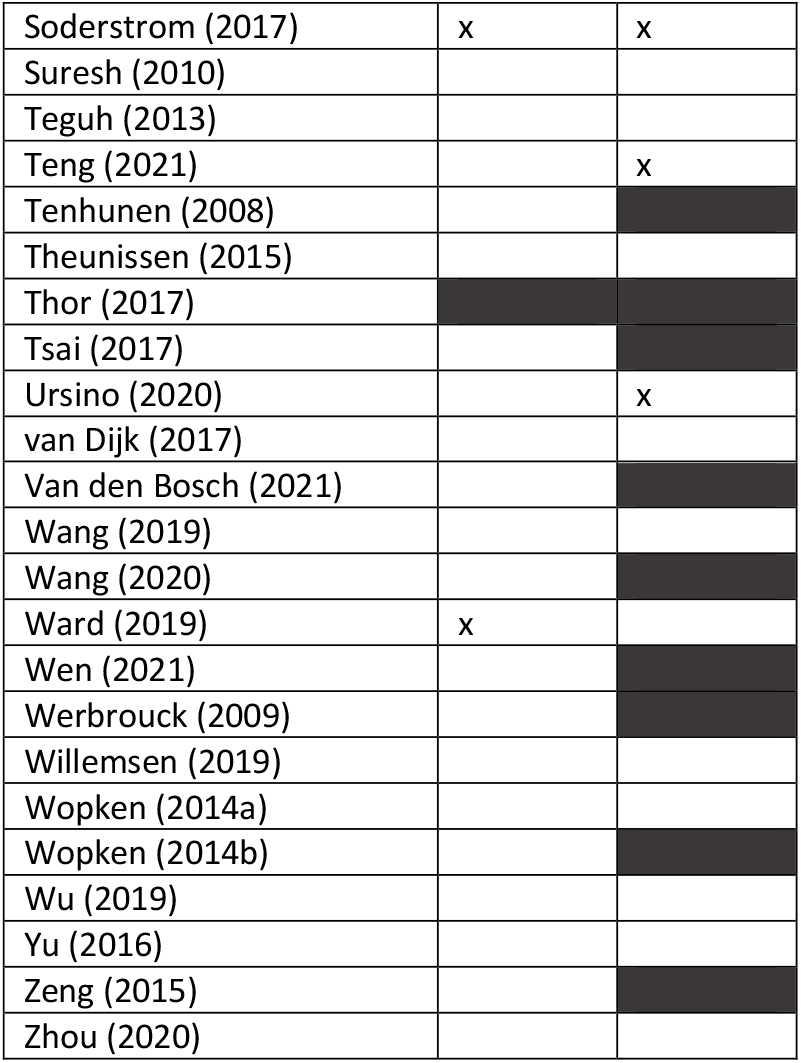
table showing the purpose and claims the studies made. Coloured spaces represent studies with a causal aim or that make causal claims, blank spaces represent studies that have a purely predictive aim or no causal claims, spaces marked with an x represent studies that are unclear.

**Appendix 3:**
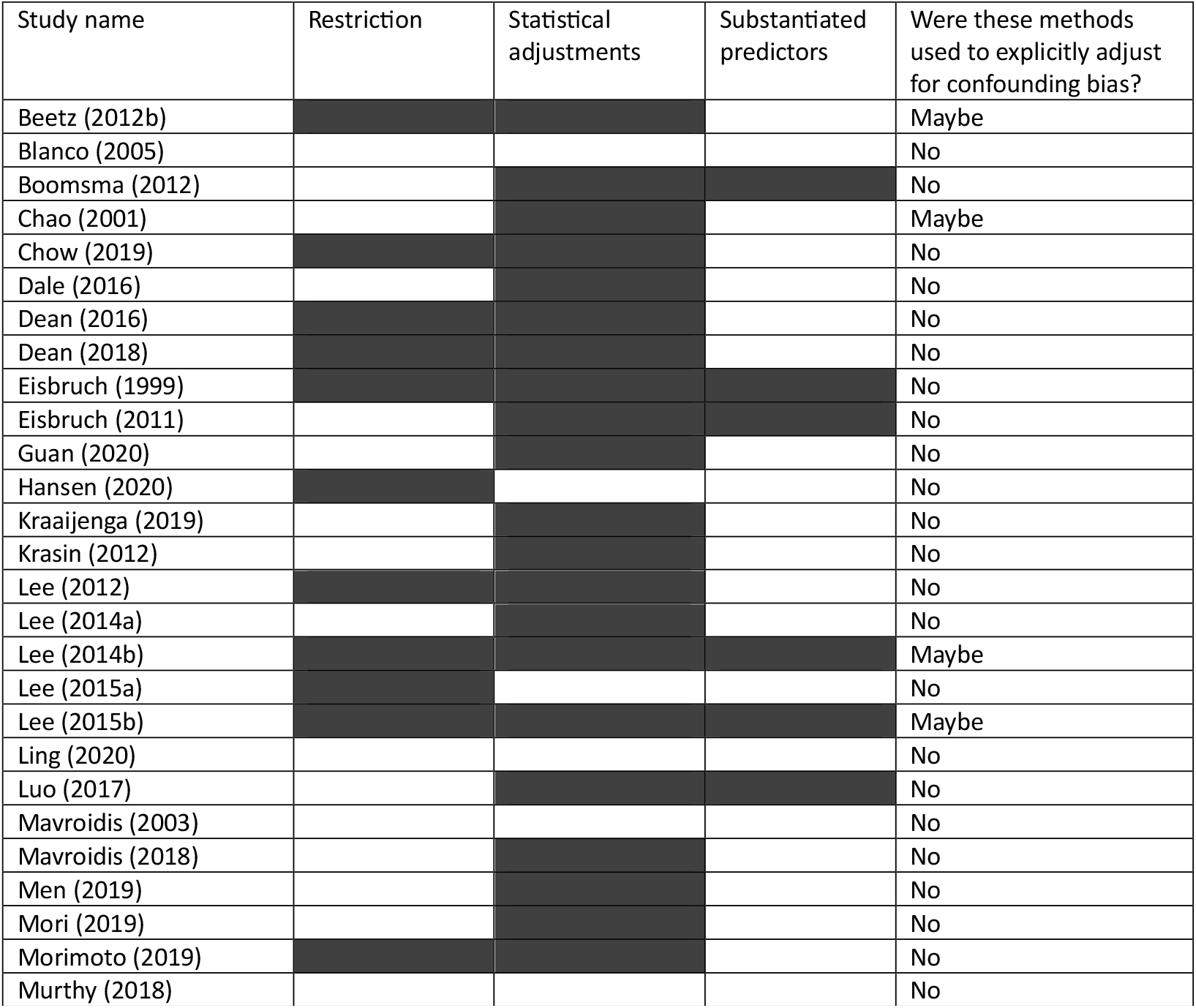
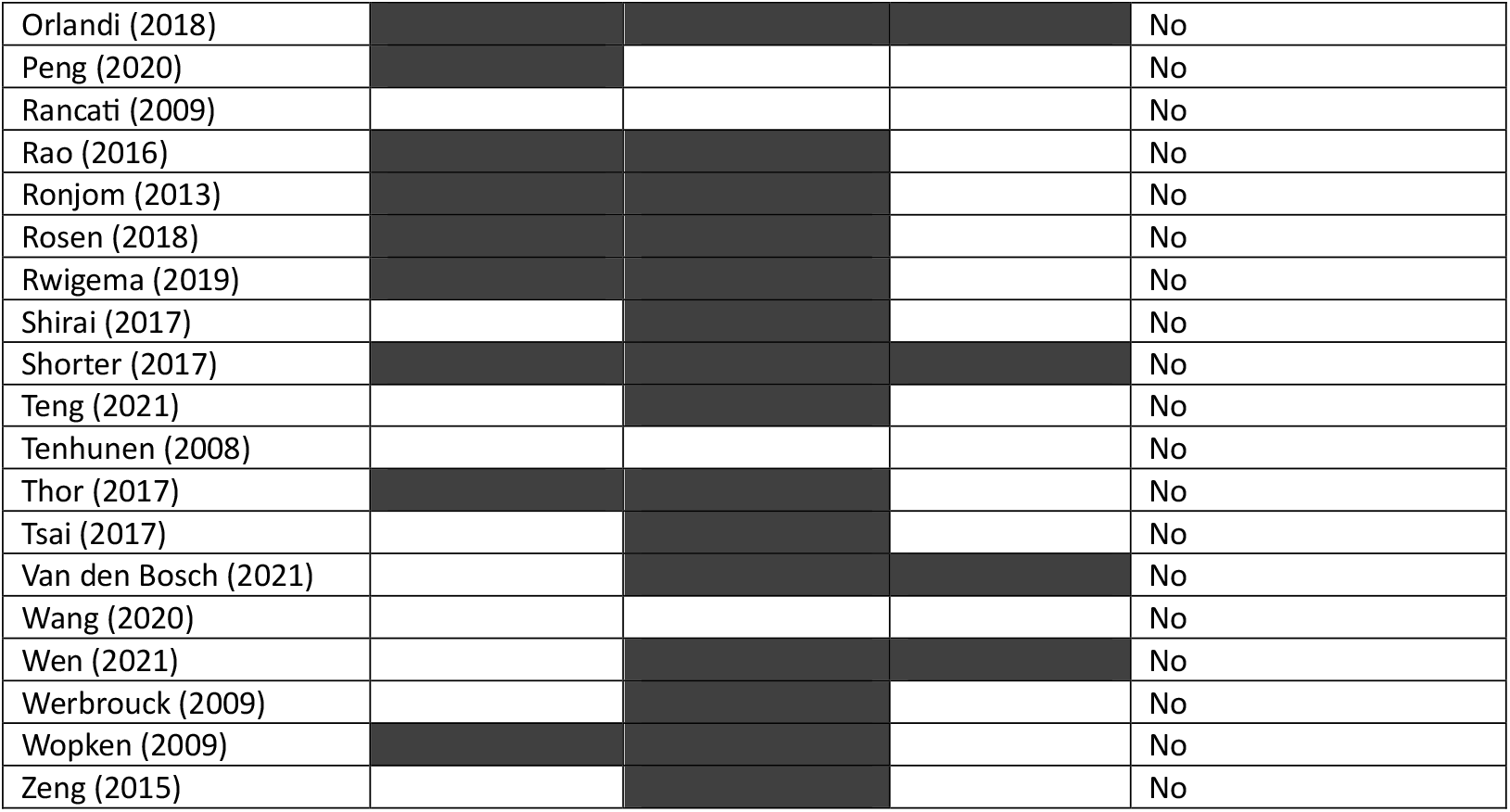

*Appendix 4: evaluation of the consistency of causal statements in existing NTCP model development studies discussing the relationship between radiation and complications in patients with lung cancer.*

We used the methods described in this paper to evaluate studies from two systematic reviews. The first review by Rodrigues et al. included clinical studies assessing the relationship between dose–volume histogram parameters and the rate of radiation pneumonitis in patients with radically treated lung cancer.^1^ There were six studies that had a causal interpretation of the results and only one study with a causal aim. This underlines our finding that researchers’ interpretations are misaligned with their intent.

The second review by Wang et al. summarized existing prognostic prediction models of non-small cell lung cancer. Only studies which were externally validated were included.^2^ Interestingly, there were no studies that had explicit causal aims or that made explicit causal claims. This could be explained by the fact that all of the studies were validated, which means that extra time and research were involved in the development of the models.

The full overview of the studies from these reviews and their classification can be found in table 6.

**Table 6:**
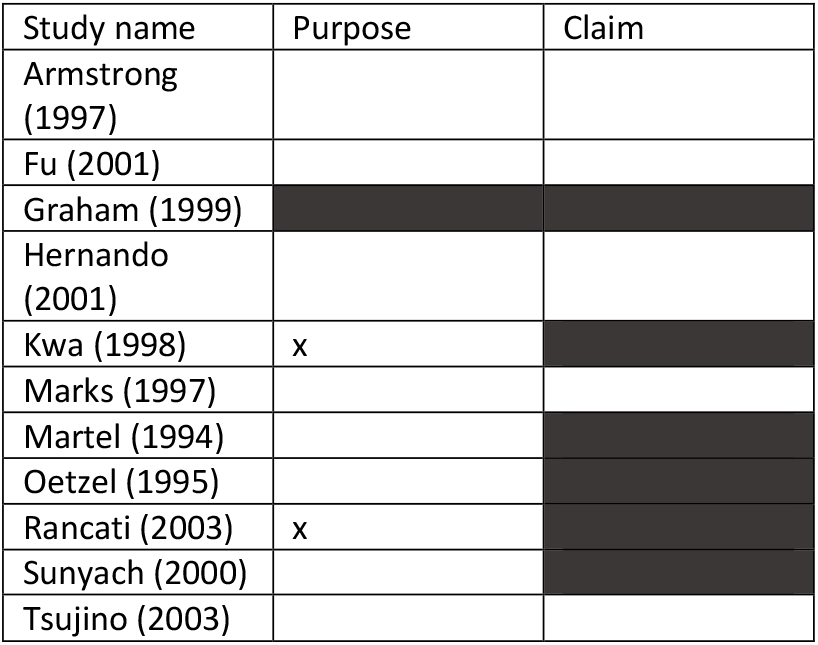
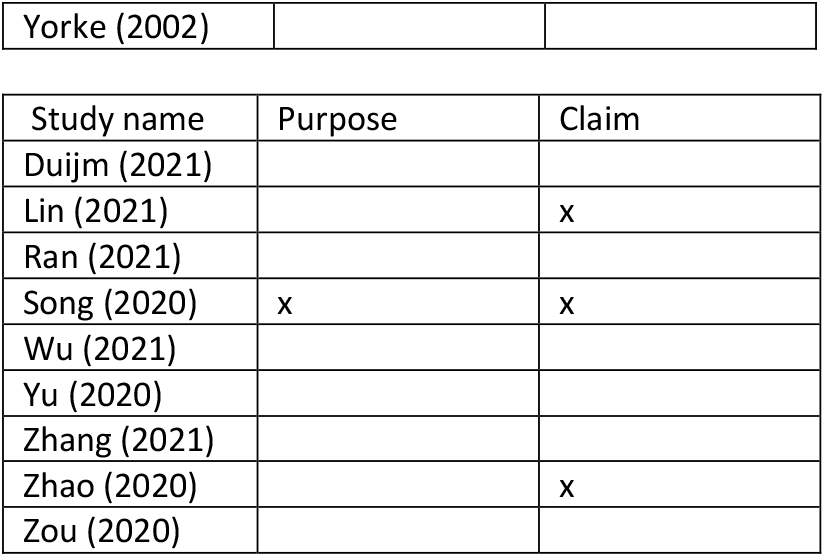
classification of studies from Rodrigues et al. (a) and Wang et al. (b). Spaces that are coloured in represent studies with a causal aim or that make causal claims, spaces that are left blank represent studies that have a purely predictive claim.

